# Persistent racial disparities in deep brain stimulation for Parkinson’s disease

**DOI:** 10.1101/2022.04.05.22273075

**Authors:** Samuel W. Cramer, Truong H. Do, Elise F. Palzer, Anant Naik, Abigail L. Rice, Savannah G. Novy, Jacob T. Hanson, Amber N. Piazza, Madeleine A. Howard, Jared D. Huling, Clark C. Chen, Robert A. McGovern

**Author notes:** **Corresponding author** Robert A. McGovern, MD, Department of Neurosurgery, University of Minnesota, 420 Delaware St SE, Mayo D429, MMC 96, Minneapolis, MN 55455.

## Abstract

We sought to determine whether racial and socio-economic disparities in the utilization of deep brain stimulation (DBS) for Parkinson’s disease (PD) have improved over time. We examined DBS utilization and analyzed factors associated with placement of DBS. The odds of DBS placement increased across the study period while White PD patients were 5 times more likely than Black patients to undergo DBS. Individuals, regardless of racial background, with two or more comorbidities were 14 times less likely to undergo DBS. Privately insured patients were 1.6 times more likely to undergo DBS. Despite increasing DBS utilization, significant disparities persist in access to DBS.

## Introduction

Surgical treatment of Parkinson’s disease (PD) with deep brain stimulation (DBS) has demonstrated efficacy that complements pharmacotherapies in managing the chronic motor symptoms of PD while improving patients’ quality of life and ability to perform daily activities. Multiple randomized clinical trials demonstrate superiority of DBS compared to medical management alone in selected PD patients.^1^ Therefore, evidence supports the efficacy and safety of DBS for PD.

Despite wide acceptance as an efficacious therapy for PD, significant disparities in DBS access have been identified, particularly for Black patients.^2,3^ In the ten years after FDA approval of DBS, Black PD patients were 5-8 times less likely to receive DBS than White patients.^2^ Since that time, much effort has been put into reducing health disparities.^4^ In this study, we examined the National Inpatient Sample (NIS) from 2002 - 2018 to determine whether disparities in DBS access has improved in the last decade.

## Methods

### Sampling

We examined NIS data from the Healthcare Cost and Utilization Project (HCUP) and the Agency for Healthcare Research and Quality.^5^ The database was queried from 2002 - 2018 using the International Disease Classification (ICD) 9 and 10 diagnostic and procedural codes (see **Figure 1A**). We identified hospitalized PD patients ≥ 18 years of age and the subset of patients admitted for DBS while excluding individuals with other neurodegenerative diagnoses.

**Figure 1:**
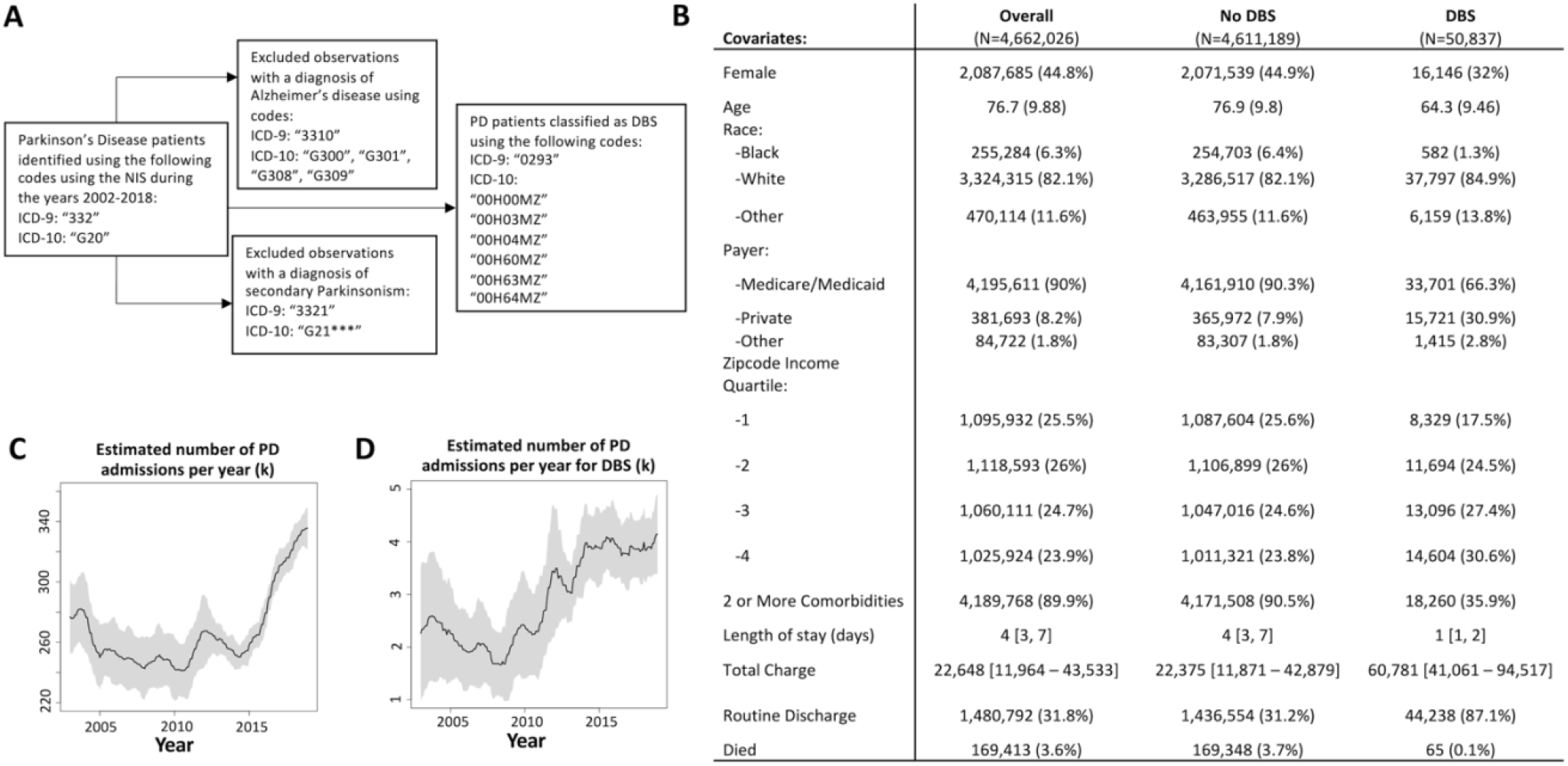
ICD codes used to define PD cohort and DBS cases (A). National estimates of patient characteristics summed or averaged across 2002 through 2018 (B). Patient characteristics are summarized by estimated mean (sample SD), median [IQR], or estimated N (estimated %) for continuous normal, continuous skewed, and categorical variables, respectively. Note that White patients predominated among both PD patients without (82.1%) and with DBS (84.9%). Black patients composed 6.4% of PD admissions not undergoing DBS and 1.3% of those admitted for DBS placement. National estimates of PD admissions (all PD admission, including for DBS) over time (C) and national estimates of DBS procedures over time (D) in thousands.

### Patient classification

The Elixhauser comorbidity software, supplied by HCUP, was used to identify comorbidities among our patient sample.^6^ Patients were dichotomized based on the number of comorbidities using a cutoff of ≥ 2 based on an accepted threshold for defining multimorbidity^7^, readmission, and surgical complications. Patients were classified as follows: PD without DBS versus PD with DBS, and number of comorbidities (< 2 versus ≥ 2 comorbidities). Patient demographics included sex, age, race, payer, and income quartile classification of patients’ zip code derived from Claritas. Based on the NIS database structure, racial categories were defined as White, Black, and Other.

### Statistical methods

All analyses were performed under the survey framework using sampling weights, clusters, and strata provided by the NIS database. Plots of procedures over time were constructed by estimating 12-month running averages with a 95% confidence band. A multivariable survey logistic model assessed the odds of a PD admission undergoing DBS with and without an interaction term for year and race. A post-hoc chi-squared test examined the association between insurance type and year stratified by race. Analyses were performed using SAS software version 9.4 or R version 3.5.1 using the “survey” package.^6^ Estimates of admission and surgical volume based on primary insurance type are displayed per 100k of the United States (US) population with that respective insurance type while those in the “Other” category are shown per 100k uninsured individuals using data from the US census bureau.^8,9^

## Results

### Characteristics of the study cohort and time trend analysis of DBS utilization

**Figure 1B** highlights characteristics of the overall cohort of PD patients, individuals not undergoing DBS, and those hospitalized for DBS placement. **Figures 1C** and **1D** illustrate the estimated number of hospitalizations for PD and patients undergoing DBS, respectively. Estimated DBS utilization increased by 82%, from ∼ 2,200 DBS surgeries in 2002 to ∼ 4,000 in 2018. Hospital admission of PD patients increased from 285,000 in 2002 to 336,000 in 2018. To quantitatively assess the apparent increase in DBS after 2010, we divided the study period into four epochs. After adjusting for ≥ 2 comorbidities, age, sex, insurance, and income, stratified by year and race, the odds of PD patients receiving DBS were comparable between the 2002 - 2005 and 2006 - 2009 period (OR 0.83, CI 0.59-1.18). However, PD patients were more likely to receive DBS during the time periods 2010 - 2013 (OR 1.45, CI 1.07-1.97) and 2014 - 2018 (OR 1.71, CI 1.25-2.34) relative to 2002 - 2005.

### Time trends in DBS placement as a function of insurer type and factors associated with DBS

**Figures 2A, 2B**, and **2C** depict patients undergoing DBS between 2002 - 2018 as a function of primary insurer type. While increased DBS utilization is observed for patients insured by Medicare/Medicaid (**Figure 2A**), private insurer (**Figure 2B**), and other insurers (**Figure 2C**), growth in DBS is most evident in Medicare/Medicaid patients as these patients account for the bulk of patients undergoing DBS.

**Figure 2:**
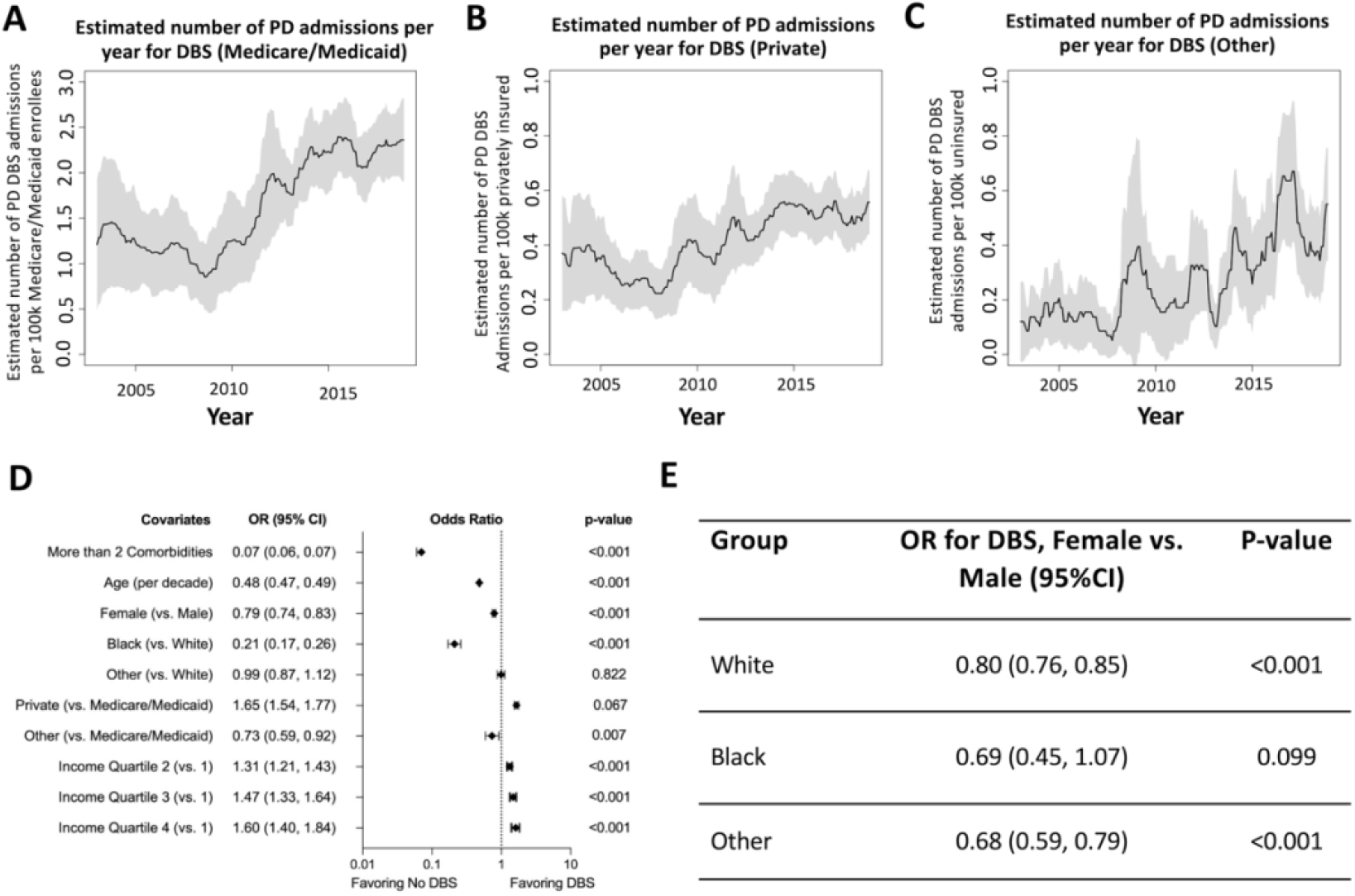
Changes in primary insurance for Medicare/Medicaid (A), Private (B) and Other (C) over time in patients undergoing DBS. For A-C, data are displayed per 100k of the overall US population with that respective insurance type while those in the “Other” category are shown per 100k uninsured individuals over the study period. Note that quantitatively, these time trends are characterized by slopes per year of 2.39, 0.08, and 0.77 for Medicare/Medicaid, private, and other insurers, respectively. Results to a multi-variable survey logistic regression models assessing the odds of DBS (D) adjusted for the year. Note that “Other” types of insurance include patients that self-pay; those that incur no hospital charges; or those that have their hospital admission paid via worker’s compensation, Title V, or other government programs. Effect of sex on DBS by race after adjusting for year, 2 or more comorbidities, age, payer, and Zip code income quartile (E).

Private insurance was associated with increased odds of DBS compared to patients insured by Medicare/Medicaid (OR 1.64, CI 1.53-1.76). Patients primarily insured in the “Other” category were less likely to undergo DBS (OR 0.70, CI 0.56-0.88). Black patients were less likely to undergo DBS than White patients (OR 0.21, CI 0.17-0.27). “Other” racial groups had similar odds of undergoing DBS compared to White patients (OR 0.99, CI 0.87-1.12). We found that females were significantly less likely to undergo DBS than males (OR 0.79, CI 0.75-0.83). Older age was associated with decreased likelihood of undergoing DBS (OR 0.48 per decade, CI 0.47-0.49). Finally, patients with ≥ 2 comorbidities were less likely to undergo DBS (OR 0.07, CI 0.06-0.07). Each subsequent income quartile was associated with increasing odds of undergoing DBS compared to the lowest quartile (all p < 0.001; **Figure 2D**).

To further evaluate the observed sex disparity in the likelihood of receiving DBS between female and male PD patients, we performed a multivariate analysis across sex based on racial background. White female PD patients were less likely than White males to receive DBS (OR 0.80, CI 0.76-0.85), while female PD patients from Other racial backgrounds also demonstrated a reduced likelihood of undergoing DBS compared to male patients from Other racial backgrounds (OR 0.68, CI 0.59-0.79). We observed a similar trend for Black female PD patients compared to Black male patients (OR 0.69, CI 0.45-1.07), though this was not significant.

Given the large observed influence of comorbidities on the likelihood of receiving DBS, we evaluated the percentage of the overall cohort of PD patients with ≥ 2 comorbidities, stratified by race. We found that among PD patients, 89.9% of White patients, 94.1% of Black patients, and 90.4% of Other patients had ≥ 2 comorbidities. We then assessed the effect of racial background on the likelihood of receiving DBS based on comorbidities and found that among Black patients with ≥ 2 comorbidities, the odds of receiving DBS was 0.19 (CI 0.14, 0.25) compared to White patients with ≥ 2 comorbidities. The odds of Black patients with < 2 comorbidities receiving DBS was 0.23 (CI 0.17, 0.30) compared to White patients with < 2 comorbidities. When comparing the odds of Black PD patients receiving DBS compared to White patients (regardless of the number of comorbidities among the racial groups) we found no significant difference in the interaction (p = 0.268), suggesting that comorbidities do not underlie the racial disparity between the likelihood of Black PD patients receiving DBS compared to White PD patients. We also compared the odds of PD patients from Other racial backgrounds to White patients and found no statistical differences between those with ≥ 2 comorbidities (OR 0.96, CI 0.82, 1.11) and those with < 2 comorbidities (OR 1.01, CI 0.88, 1.16). Finally, among PD patients from Other racial groups (regardless of the number of comorbidities), we found no significant difference in the interaction (p = 0.495).

### Disparity in DBS utilization rate over time

Prior work has shown Black PD patients in the US are 5-8-fold less likely to undergo DBS relative to White patients.^2,3^ Correction of this disparity would require that the likelihood of DBS utilization in Black patients outpace that of White patients by 5-8 to 1. We tested whether this occurred by comparing the odds of DBS placement in Black and White patients pre- and post-2010 (**Figure 4A**). The odds of undergoing DBS during the period 2010-2018 versus the period 2002-2009 for Black PD patients was 2.24 (CI 1.20-4.19). For the White patients, the corresponding odds ratio was 1.76, (CI 1.38-2.24). A direct comparison of Black and White patients undergoing DBS in 2002 - 2009 demonstrates that the odds of Black patients receiving DBS was 0.17 compared to Whites after adjusting for confounders (OR 0.17, CI 0.10-0.29, p < 0.001). In 2010 - 2018, the odds of Black patients receiving DBS was 0.21 compared to Whites (OR 0.22, CI 0.17-0.28, p < 0.001; **Figure 4B**). Therefore, more Black and White patients have received DBS since 2010, but the racial disparity in DBS has not meaningfully changed. There was no significant change in insurance type between racial groups (**Figure 4C**).

## Discussion

As the US population ages, the prevalence of PD has been increasing^10^ as reflected in the overall increase in hospital admissions of these patients in the last two decades (**Figure 1C**). Accordingly, and perhaps in concert with increased acceptance of DBS over time, there has been significant growth in DBS in PD patients during the same period (**Figure 1D**). Growth in DBS, however, has not meaningfully changed the racial disparity in DBS utilization over the last decade. After controlling for factors associated with DBS placement, including age, comorbidities, insurance type and income quartile, Black patients were five times less likely to undergo DBS than White patients in the last decade, almost identical to the ratio seen in the prior decade (**Figure 4B**). Analysis of the likelihood of receiving DBS across racial groups using a finer timescale (3-year increments across the study period) did not demonstrate a significant change across the study period (data not shown), suggesting that the racial disparities between have not narrowed with time. The findings demonstrate persistent disparities in the clinical practice of DBS that warrant consideration for improvement.

As an elective surgery aimed at improving the quality of life in PD patients, surgeons and DBS teams must minimize the most common complications of DBS surgery (hemorrhage, infection) in order to justify its benefits. The selection process of DBS candidates is one major aspect of this risk management and it is therefore not surprising that PD patients with ≥ 2 comorbidities are less likely to undergo surgery. We and others have previously demonstrated that selection criteria for DBS patients appeared to be getting more liberal over time with surgical patients getting increasingly older and more likely to have comorbidities.^11^ We considered that differences in the number of comorbidities may affect the likelihood of receiving DBS and that these differences may be different between Black and White PD patients. We tested this hypothesis by comparing the likelihood of Black patients receiving among those with < 2 comorbidities and those with ≥ 2 comorbidities and found that Black patients from both groups were approximately 5 times less likely than White patients to receive DBS (**Figure 3**). Furthermore, there was no significant difference in the interaction among the comparison between these two groups which suggests that the number of comorbidities does not contribute to the observed racial disparity. Although previous studies have demonstrated racial differences in the likelihood of comorbidities such as obesity and diabetes mellitus^12^, this does not appear to be a significant factor in the racial disparity between Black and White PD patients who receive DBS. One limitation of this analysis is that it only addresses the quantity of comorbidities and not necessarily the severity or quality of comorbidities. Nevertheless, the presence of two or more comorbidities remains the strongest predictor of not undergoing DBS and demonstrates that PD patients still undergo a rigorous selection process.

**Figure 3:**
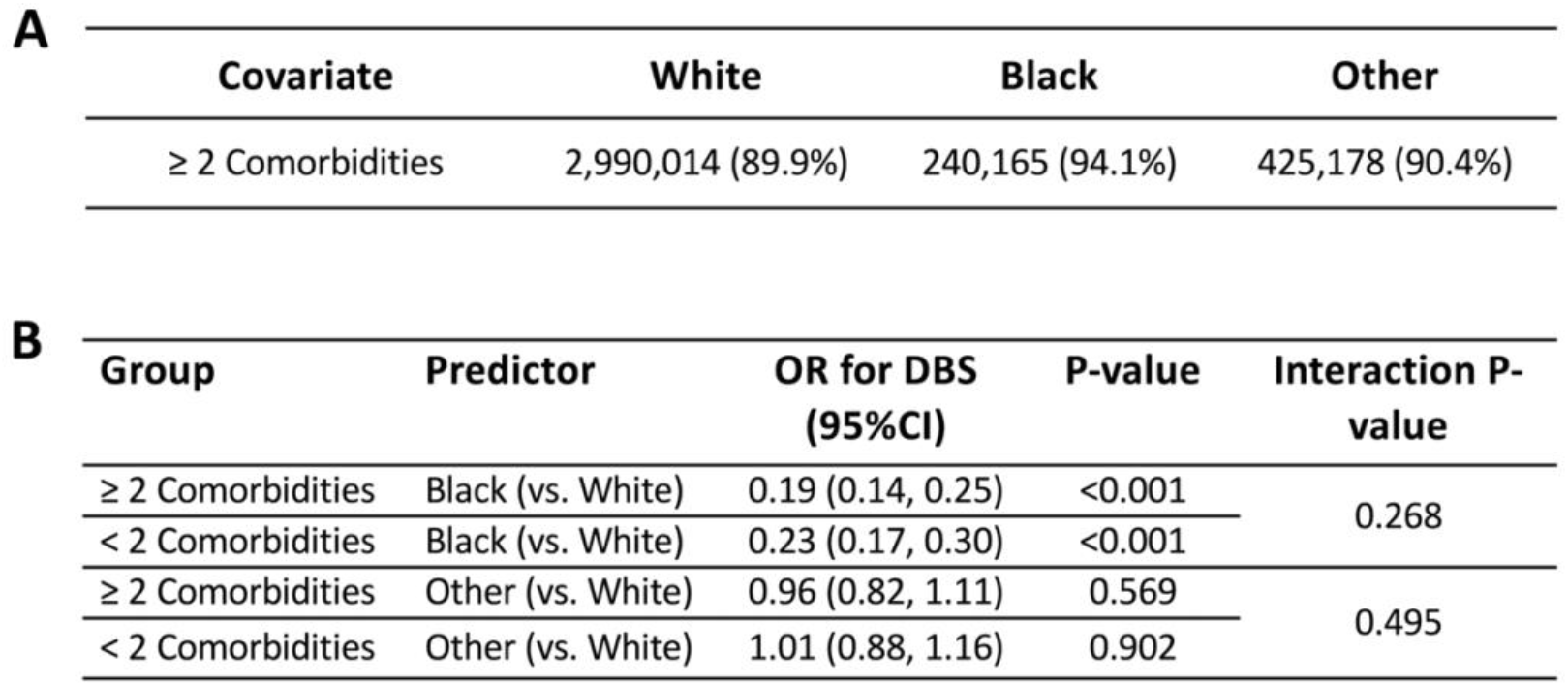
Descriptive statistics showing comorbidity by race, N (%), (A). Assessment of the effect of race on likelihood of DBS by comorbidities after adjusting for year, sex, age, payer, and Zip code income quartile (B).

**Figure 4:**
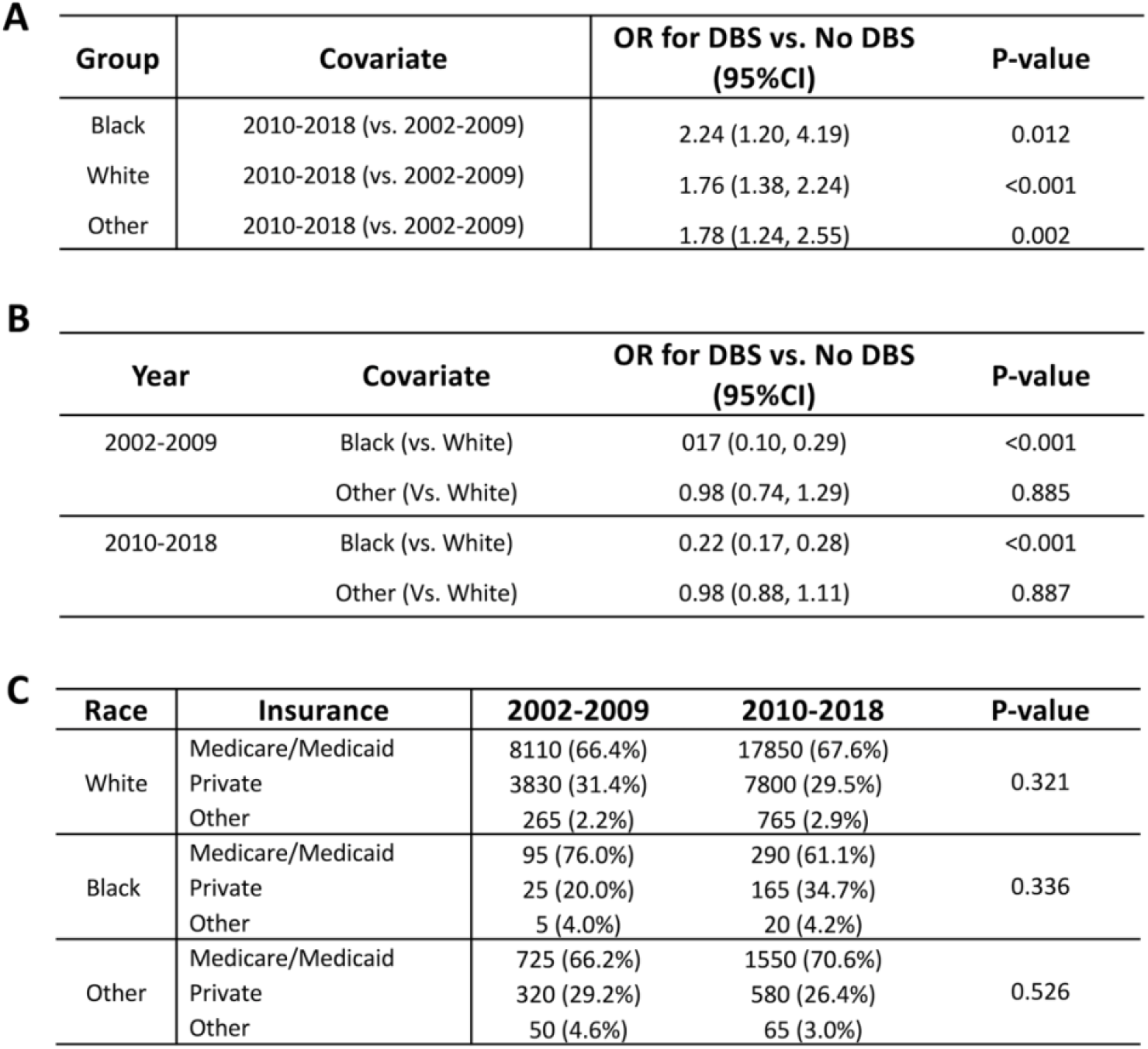
Results of a multivariable survey logistic regression model assess the association between race and DBS over time for Black patients, White patients, and Other (A). Direct comparison of odds of undergoing DBS based on race across years (B), note that the model adjusted for 2 or more Elixhauser comorbidities, age, sex, insurance type, and zip code income quartile. Post-hoc survey chi-squared analysis to test if insurance type for those who received DBS differs across the study period by race (C).

Differences in surgical outcomes across racial groups may also contribute to the racial disparity in DBS utilization. For example, if White patients tend to have better outcomes than Black patients or patients from Other racial groups, this may bias the recommendation for DBS therapy for the group with better outcomes. Unfortunately, there is limited literature to draw upon to address this question as it pertains to possible racial differences in postoperative outcomes following DBS surgery. One recent study found that White and non-white PD patients were similarly likely to be routinely discharged following DBS surgery.^13^ However, there is evidence that postoperative outcomes (including length of stay and complications) are worse for Black patients compared to White patients following a range of major surgeries.^14–18^ Black patients also have higher postoperative mortality rates.^19–21^ Therefore, to what extent surgeon/institutional experience with racial disparities in outcome following DBS surgery may play a role in patient selection is unclear and should be explored in future analyses examining factors underlying the disparities in DBS for PD.

The sex disparity in the utilization of DBS for PD is another notable finding. We observed that female PD patients are less likely to receive DBS than male patients. Further analysis among genders across racial backgrounds demonstrates that while the odds of White females receiving DBS is 0.8 compared to white males, females from Other racial backgrounds are even less likely to receive DBS compared to males from Other racial backgrounds. This trend was also observed for Black females compared to Black males though the difference was not statistically significant, likely owing to inadequate power to detect a difference. The largest group of PD patients in the US is White males (gender prevalence estimated at 1.55 male: 1. female).^22^ Despite the increased prevalence of PD in males, the number of patients referred for DBS is disproportional to the prevalence data.^23,24^ A previous retrospective study at a single institution found that 75% of PD patients referred for DBS were male. Furthermore, among patients that were referred and chose not to undergo DBS, patient preference was the dominant reason cited by female patients for not pursuing DBS, which was significantly more common than among male patients.^22^ Therefore, sex referral bias as well as patient self-selection may in part underlie the observed sex disparities, factors which may be amplified in non-White PD patients.

Increased DBS utilization has not yielded uniform benefit across racial groups. Epidemiological studies examining racial differences in incidence and prevalence of PD have demonstrated mixed findings.^25^ Absent racial epidemiological differences, the persistent disparity that Black PD patients are five-fold less likely to undergo DBS relative to White patients is notable. It should also be noted that the disparities in the utilization of DBS are unfortunately not unique to DBS surgery. Multiple studies have demonstrated that Black patients as well as patients from other racial minorities are less likely to receive surgery among a range of different procedures.^26,27^ Furthermore, in the case of epilepsy patients undergoing surgery over a similar time period as this study, the odds ratio for Black patients receiving surgery was approximately 0.5 compared to White patients.^28^ Our finding that the odds of Black patients receiving DBS is 0.2 compared to White patients is among the lowest for all elective surgery types. This highlights the importance of systemic factors inherent in the US medical system that contribute to the pervasive inequities of surgical care across racial groups. Several factors may underlie this disparity including unconscious/implicit or conscious/explicit bias which contributes to underrepresentation of Black patients in specialty clinic referrals.^2,29^ Interventions to change physician behavior generally focus on improving adherence to evidence-based guidelines but these practices could be adapted to improving disparities in referrals or selection for DBS. Active methods including continuing education, computerized decision support systems/reminders and financial incentives that have been shown to effectively modify behavior.^30^ Thus, while documenting disparities in healthcare increases awareness, reducing disparities depends upon changing physician behavior on a local, institutional level beginning with medical training.^31^ For example, training physicians to utilize strategies including individuation^32^, perspective taking^33^ and other strategies^31^ to combat implicit bias. Incorporating these strategies into undergraduate medical education would further spur systemic change necessary to truly reduce disparities in medical care.

While changing individual behavior (either physician or patient) has been a main focus in disparities research, recent studies have highlighted that many additional factors such as structural barriers should be considered as contributing to racial disparities. For example, a potential barrier to access surgical care of PD is one of geography. Regional differences in the density of neurologists and neurosurgeons across urban and rural settings may influence access to care. Despite this hypothetical concern, a previous study examined this question and demonstrated that this does not appear to be a significant factor.^2^ While *density* of specialty care physicians and surgeons may not be a factor, *access* to specialty care via referral pattern biases or other mechanisms may underlie some of these disparities. Future multicenter registries that examine detailed patient demographic information (which is not available in a national dataset) would be helpful to better delineate the role geography may play in access to DBS therapy.

Socioeconomic status, and its complex association with many other factors including racial background, access to transportation, social support, and time off of work for surgery and subsequent follow-up visits (an important consideration given that frequent clinic visits are required for optimizing DBS therapy), represents another factor that may contribute to racial inequities in access to DBS. In agreement with a prior study,^3^ we found that increasing income by zip-code was a strong predictor of increased likelihood of receiving DBS compared to the lowest income quartile. In fact, at 1 year follow up of DBS placement, PD patients with higher household incomes achieve better functional outcomes,^34^ a factor which may bias referrals for DBS therapy toward higher income patients. Conversely, prior investigations suggest that Medicare coverage (a surrogate for lower socioeconomic status) amongst Black PD patients may lead to a DBS surgery non-use decision.^2^ Significant differences in the medical management of PD have also been reported based on race and socioeconomic status.^35^ Among surgical care more broadly, there is evidence to suggest that lower socioeconomic status is associated with reduced likelihood of receiving surgical care.^36–38^ Socioeconomic status, therefore, is an important factor in the likelihood of receiving DBS therapy.

Marketing of DBS therapy may play a role in racial disparities in the utilization of DBS for PD. As one of the presumed drivers of DBS growth over the past two decades, marketing efforts may not be uniformly received and/or effective across racial backgrounds. The effects of device marketing for DBS systems have not been specifically investigated but the effects of direct to consumer marketing of pharmaceuticals has been studied. For example, Black patients are less likely to be exposed to direct to consumer marketing of pharmaceuticals compared to White patients while Black patients are more likely to be influenced by such efforts.^39^ Patient preference for innovative technologies including implantable medical devices may differ along racial lines which could play a role in how marketing efforts are received, although a previous study demonstrated that Black and White patients have similar preferences for implantable medical devices.^40^

Patient-specific factors may also contribute to racial disparity. Black PD patients under-report motor impairment compared to White patients and therefore present later in the disease process, delaying their diagnosis and treatment.^25^ Studies examining this phenomenon have demonstrated that Black patients are more likely to view PD as a part of normal aging and therefore may be less likely to act on their symptoms to seek treatment.^41^ A large single-center study examining racial disparities in PD among 1159 patients showed greater disability and disease severity in Black patients.^35^ This finding is supported by Weuve et al. who found that Black people may have higher rates of cognitive impairments, dementia, or other comorbidities among aged patients with Parkinson’s disease which may differentially limit their candidacy for DBS surgery,^42^ although we found no differences in co-morbidities among racial groups. Medical mistrust among Black patients due to repeated past mistreatment in studies such as the Tuskegee Syphilis Study^43^ is also a factor contributing to delays in diagnosis and treatment. One recent study identified perceived discrimination due to income or insurance type as factors most strongly associated with medical mistrust among Black patients with perceived discrimination due to race/ethnic background more weakly associated.^44^ Highlighting patient factors that may influence referral or selection for DBS therapy is important. This is not because the patient’s individual behavior fundamentally reduces the likelihood of receiving DBS but rather that the treating or referring physician must recognize patient-specific factors that may unfairly bias the physician’s treatment recommendations.

As discussed above, the exact causes of the persistent racial disparities in the use of DBS for PD are complex and multifactorial. Important open questions remain regarding DBS therapy. What is the rate that DBS is offered across racial groups? What is the rate at which DBS is offered but declined across racial groups? While we cannot investigate these questions with our current data set, answering these questions may provide some degree of clarity regarding the root cause of racial disparities in DBS therapy for PD. For example, multicenter registries or single institution studies which track referrals and reasons for acceptance/decline would be able to provide some insight into the specific reasons for racial disparities and highlight potential avenues for improvement.

Increasing utilization of DBS for PD has not improved racial disparities in access to this important therapy. While previous studies have identified racial disparities in DBS therapy, we believe that it is important to point out that nearly a decade after these findings were first reported, racial inequities in DBS therapy remain essentially unchanged. Highlighting persistent racial disparities in DBS is a preliminary effort to reform US healthcare delivery to ensure equal access to life altering treatments regardless of racial background.

## Data Availability

All data produced in the present study are available upon reasonable request to the authors

https://www.hcup-us.ahrq.gov/tech_assist/dua.jsp

## Authorship

SWC, THD, CCC and RAM contributed to the conception and design of the study. SWC, THD, CCC, RAM, EFP and JDH were responsible for acquisition and analysis of data. SWC, THD, EFP, AN, ALR, SGN, JTH, ANP, MAH, JDH, CCC, and RAM contributed to drafting the text and preparing the figures.

## Ethics statement

Approval to utilize the HCUP NIS database was obtained after registration with HCUP. In accordance with The Health Insurance Portability and Accountability Act of 1996, use of the data available via HCUP does not require review by an institutional review board (IRB) is not required because it is a limited data set. All necessary patient/participant consent has been obtained and the appropriate institutional forms have been archived.

## Acknowledgements

*This study was supported in part by funding from MnDRIVE, a collaboration between the University of Minnesota and the State of Minnesota. The authors declare that they have no conflict of interest*.

